# SARS-CoV-2 Seroprevalence and Risk Factors among Convalescents in Sichuan Province, China: a retrospective cohort study

**DOI:** 10.1101/2021.05.04.21256355

**Authors:** Lijun Zhou, Cheng Li, Huiping Yang, Heng Yuan, Ming Pan, Xiuwei Cheng, Chongkun Xiao, Xiaoyan Su, Yuanfang Zhu, Jianan Xu, Jianxiang Tang, Xunbo Du, Huanwen Peng, Chen Xiao, Tao Huang, Hongxiu Liao, Deqiang Xian, Hongxiu Liao, HaoZhou Wang, Wenwu Liu, Zhou Ping, Zhengdong Zhang, Liu Juan, Xianping Wu, Tao Zhang

**Author notes:** Lijun Zhou, Cheng Li and Huiping Yang contributed equally to this maniscript. Xianping Wu and Tao Zhang contributed equally to this maniscript. **Correspondence:** Tao Zhang, Department of Epidemiology and Health Statistics, West China School of Public Health and West China Fourth Hospital, Sichuan University, Renmin South Road 3rd Section NO.16, Chengdu 610041, Sichuan.

## Abstract

**Objectives:** To understand SARS-CoV-2 seroprevalence of convalescents and assess their the immunity. Furthermore, we intend to explore the association between antibody levels and with demographic factors.

**Methods:** 177 COVID-19 convalescents in Sichuan Province were voluntarily participated in our study. 363 serum samples were collected from June, 2020 to November, 2020. Duration of seroprevelance in these convalescents and their demographic characteristics were described, and the risk factors to antibody levels were analysed.

**Results:** Men had more than twice the odds of having IgM antibody positive compared with women (OR=2.419, 95% CI:[1.232, 4.751]). Participants without symptoms were nearly 0.5 times IgG seropositive than those with symptoms (OR=0.455, 95% CI: [0.220, 0.940]). People aged≥60 years were nearly 3 times IgG seropositive than those who aged < 20 years (OR=2.986, 95% CI: [1.058, 8.432]). Seroprevalence in asymptomatic declined quicker than symptomatic.

**Conclusions:** Age and gender may affect the antibody levels and seroprevalence. Asymptomatic appeared more easier to turn to seronegative than symptomatic.

## Introduction

The global pandemic of coronavirus disease 2019(COVID-19), an emerging infectious disease resulted from severe acute respiratory syndrome coronavirus 2 (SARS-CoV-2), has pose enormous threat to public health[1]. The epidemiological and serological characteristics of patients with COVID-19, have been explicitly reported[2], but the seropervalence, serological characteristics and immunnity of convalescents remain largely obscure.

Antibody response is crucial to eliminate viral infection[3], and the seroprevalence of specific serum antibodies immunoglobulin M (IgM) and immunoglobulin G(IgG) against SARS-CoV-2 can provide immune protection. Understanding seroprevalence dynamic of SARS-CoV-2 assists to assess the immune levels of convalescents, and it also helps to predict the potential protection in the future [4]. As the number of patients recovering from the SARS-CoV-2 continues to increase, the duration of individual serological responses has also sparked public attention[5]. It is important to know whether and how demographic factors (such as age, gender, origin and type) affect serological responses during SARS-CoV-2 infection. In several countries, initial case reports have symptomatic to be re-infected with SARS-CoV-2 [6]. There is an urgent need to understand how long the antibody response to the virus will last after infection. For one thing, monitoring specific antibodies can benefit to distinguish vaccine-related antibodies from infected-stimulated ones[7]. For another, the study of serological dynamic also help to understand the associated serological epidemiology.

IgM is the first immune response against viral infection, and IgG production lags behind IgM but provides long-term immunity[8], similar to coronavirus infections such as SARS and MERS[9]. Usually IgM antibodies last less time in the body, and previous studies in SARS patients have shown that specific IgM antibodies last for 13 weeks in the body[10], as well as IgG antibodies are longstanding with an average of about 2 years[11]. The decline of IgM is seen as a indicator of virus clearance and convalescents could have robust immunity against re-infection with positove antibody, the reduction of IgG prompt serious concerns on the robustness and persistence of immunity after recovering[12], which is primary for developing vaccine and making immunity strategy. Studying the risk factors and dynamic change of different antibodies can help to understand the body’s resistance to SARS-CoV-2 at different stages. At the same time, information regarding the persistence of immunity to SARS-CoV-2 infection facilitate the continued development of new vaccines and clinical therapeutics[13]. In our study, the serological level of 177 convalescents of SARS-CoV-2 in Sichuan Province were reviewed and analyzed. At present, the literature reported that the seroprevalence of specific IgM antibodies and IgG antibodies persisted to 8 months[14], but the positive antibody duration still needs to update.

However, most of previous researches have mainly spotlighted on the acute response within several weeks after clinical onset in SARS-CoV-2. While the number of convalescents infected with SARS-CoV-2 started to escalate, evaluating and understanding the immune response becomes even more essential. This study intends to assess whether seroprevalence is associated with demographic factors, such as gender, age, type (symptomatic cases or asymptomatic cases) and travel history(domestic or international). Meanwhile, we aims to describe the serum dynamic changes and durability of convalescents of SARS-CoV-2.. It is expected that serological study of SARS-Cov-2 convalescents during the recovery period will benefit to understand the immunological response to SARS-CoV-2 infection. Moreover, It also provides an auxiliary scientific basis for clinical development and evaluation of SARS-CoV-2 vaccine.

## Methods

### Study design

This study is a retrospective cohort study that included 177 convalescents in Sichuan Province as of November 23, 2020. All subjects voluntarily joined serological researches with informed consent. 363 serum samples and demographic characteristics of 177 convalescents were collected.

### Data and specimen collection

The data was collected by the Centers for Disease Control and Prevention in Sichuan Province, and consisted of the demographic characteristics of the 177 convalescents and their longitudina antibody results (363 serum samples) till November 23, 2020. Specimens were collected from June 23, 2020 on the basis of voluntary informed consent of COVID-19 convalescents. Volunteers can withdraw from the study at any time in accordance with the principle of informed consent. All 363 serum samples were detected by the Institute of Microbiology and Analysis.

### Detection of IgG and IgM

Non-anticoagulant specimens (intravenous blood collection) were collected for all subjects, 3mL for children (<5 years), and 5mL for others. Serum samples were collected and loaded into sealed bags in accordance with Class A transport packaging, refrigerated and transported to the local CDC laboratory for serum separation. The isolated serum was stored in a 1.5 mL freezed deposit tube at -20 degrees C. The Mike i1000 fully automated luminescent immunoanalystator (base fluid lot number: 0520153; reagent lot number: 0520031,0520032; reaction cup lot number: 0720582;) was utilized to test serum by the principle of direct chemical luminescence immunoanalysis.

### Ethical approval

The survey was discussed with the Ethics Committee of Center for Disease Control and Prevention in Sichuan Province, who reviewed the content. And the ethical approval was given. All participants assented informed consent before participation, and this study was conducted in accordance with Good Clinical Practice(GCP). This study was performed in compliance with all relevant ethical regulations and the protocol for human subject studies was approved by the Center for Disease Control and Prevention in Sichuan Province(SCCDCIRB-2020-007).

### Statistical analysis

Descriptive statistics were utilized to summarize the demographic characteristics of the cohort and significant study outcome variables. Frequency and composition ratio were used for categorical variables, Chi–square test and Fisher’s exact test were used for comparing categorical variables. Multivariate logistic regression was used to calculate odds ratios and 95% confidence interval.

Demographic characteristics consisted of variables including gender, age groups (< 20 years old, 20-40 years old, 40-60 years old and ≥ 60 years old), travel history(domestic or international), type (symptomatic or asymptomatic) and antibody results (positive or negative). We considered the antibody results as outcome, and it was divided into two groups (positive or negative), previous studies have found that gender and age were related to the results of the antibody[15]. Frequency and composition ratio were used to describe the total number of specific antibodies of different durations. The seroprevalence changes and positive rates of specific antibodies IgM and IgG over time were plotted. All analyses were performed by Stata 16.0 software, the p-value less than 0.05 in this paper was considered statistically significant.

## Results

### Demographic and Clinical Characteristics

By November 23, 2020, A total of 177 participants (83 men; 94 women) were recruited. The median of age was 47 years (IQR = 33-57 years), asymptomatic cases(35 years) are youger than symptomatic(47 years). The types of cases were statistically different among different age groups(χ^2^= 14.0671,*P*=0.003).The descriptive analysis of 177 convalescents with SARS-CoV-2 infections were included (Table 1). For serology results, 152 were positive and 25 were negative. Only two subjects had international travel history.

**Table 1.** Demographic and Clinical Character istics.

|  | Total | asymptomatic | symptomatic | *P* |
| --- | --- | --- | --- | --- |
|  | No | No. (%) | No. (%) |  |
|  | 177 | 21(11.86) | 156(88.14) |  |
| Gender |  |  |  |  |
| Female | 94 | 8(8.51) | 86(91.49) | 0.142 |
| Male | 83 | 13(15.66) | 70(84.34) |  |
| Age,median(IQR) | 47(33-57) | 35(18-58) | 47(33-57) |  |
| < 20 years | 15 | 6(40.00) | 9(60.00) | 0.003 |
| 20-40 years | 53 | 6(11.32) | 47(88.68) |  |
| 40-60 years | 71 | 4(5.63) | 67(94.37) |  |
| ≥60 years | 38 | 5(13.16) | 33(86.84) |  |
| Serology assessment |  |  |  |  |
| Negative | 25 | 5(20.00) | 20(80.00) | 0.185 |
| Positive | 152 | 16(10.53) | 136(89.47) |  |
| Travel history |  |  |  |  |
| Domestic | 2 | 1(50.00) | 1(50.00) | 0.224 |
| International | 175 | 20(11.43) | 155(88.57) |  |

Values are medians (IQR) or n (%). Groups were compared using Chi-square test or Fisher's exact test.
Abbreviations: IQR, inter-quartile range

### Specific Antibodies IgM and IgG Levels

The levels of different antibodies in 177 patients infected SARS-CoV-2 were further described (Table 2). As time went by, largely of patients tended to IgG positive and IgM negative. The proportion of IgM positive in infected women was significantly lower than men, but the proportion of IgG positive was similar between different genders. People who aged ≥60 years have higher IgG (94.74%)seroprevalence than IgM(10.53%), IgG positive percentage of different age groups was statistically difference(*χ*^*2*^= 10.8514, *P*=0.013). Asymtomatic was lower than symptomatic for both IgG and IgM to SARS-CoV-2 seroprevalence.

**Table 2.** Specific Antibodies IgM and IgG Levels

|  | Total | IgM No. (%) |  | *P* | IgG No. (%) |  | *P* |
| --- | --- | --- | --- | --- | --- | --- | --- |
|  |  | negative | positive |  | negative | positive |  |
|  | 177 | 154(87.01) | 23(12.99) |  | 26(14.69) | 151(85.31) |  |
| Gender |  |  |  |  |  |  |  |
| Female | 94 | 86(91.49) | 8(8.51) | 0.059 | 11(11.7) | 83(88.3) | 0.232 |
| Male | 83 | 68(81.93) | 15(18.07) |  | 15(18.07) | 68(81.93) |  |
| Age |  |  |  |  |  |  |  |
| < 20 years | 15 | 14(93.33) | 1(6.67) | 0.376 | 5(33.33) | 10(66.67) | 0.013 |
| 20-40 years | 53 | 48(90.57) | 5(9.43) |  | 12(22.64) | 41(77.36) |  |
| 40-60 years | 71 | 58(81.69) | 13(18.31) |  | 7(9.86) | 64 (90.14) |  |
| ≥60 years | 38 | 34(89.47) | 4(10.53) |  | 2(5.26) | 36(94.74) |  |
| Type |  |  |  |  |  |  |  |
| Asymptomatic | 21 | 20(95.24) | 1(4.76) | 0.318 | 5(23.81) | 16(76.19) | 0.209 |
| Symptomatic | 156 | 134(85.9) | 22(14.1) |  | 21(13.46) | 135(86.54) |  |
| Travel history |  |  |  |  |  |  |  |
| Domestic | 2 | 1(50.00) | 1(50.00) | 0.244 | 0(0.00) | 2 (100.00) | 1.000 |
| International | 175 | 153(87.43) | 22(12.57) |  | 26(14.86) | 140 (85.14) |  |

Values are n (%). Groups were compared using Chi-square test or Fisher's exact test.
Abbreviations: IgG, immunoglobulin G; IgM, immunoglobulin M

### Multivariate Logistic Regression Analysis of Positive Antibody for IgM

The occurrence of positive IgM antibody was taken as the dependent variable, gender and type were considered as the independent variables (Table 3), female and symptomatic cases as the reference group respectively. Different age groups were dummy variables. The age group younger than 20 years was the reference group. Previous studies have found that gender and age were related to the outcome levels of antibodies[16]. Multivariate logistic regression analysis showed that age and type of infection were not related to IgM antibody results. However, after adjustment for infected type and age, men had more than twice the odds of having IgM antibody positive compared with women (OR=2.419, 95% CI:[1.232, 4.751]).

**Table 3.** Multivariate Logistic Regression Analysis of Positive Antibody for IgM

| | $\beta$ | OR | $P$ | 95% CI. | |
| --- | --- | --- | --- | --- | --- |
|  |  |  |  | lower | upper |
| Gender | 0.883 | 2.419 | 0.010 | 1.232 | 4.751 |
| Type | -1.074 | 0.342 | 0.165 | 0.075 | 1.557 |
| Age(<20 years) |  |  |  |  |  |
| Age(20-40 years) | 0.676 | 1.967 | 0.544 | 0.221 | 17.470 |
| Age(40-60years) | 1.336 | 3.806 | 0.219 | 0.452 | 32.060 |
| Age(≥60 years) | 0.713 | 2.040 | 0.525 | 0.227 | 18.320 |

Abbreviation: OR: odds ratio; CI: confidence interval ;

### Multivariate Logistic Regression Analysis of Positive Antibody for IgG

The antibody level of IgG antibody(positive or negative) was taken as the dependent variable, gender and type of infected person as the independent variables (Table 4), female and symptomatic cases were taken as the reference group. Different age groups were dummy variables. The age group younger than 20 years was the control group. While we excluded travel history due to its improper composition ratio. Multivariate logistic regression analysis showed that gender and type of infection were not related to IgG positive result. Participants without symptoms were nearly 0.5 times more likely to be seropositive than those with symptoms (OR=0.455, 95% CI: [0.220, 0.940]). People aged 60 years and older were nearly 3 times IgG seropositive than those who aged less than 20 years(OR=2.986, 95% CI: [1.058, 8.432]).

**Table 4.** Multivariate Logistic Regression Analysis of Positive Antibody for IgG

| | $\beta$ | OR | $P$ | 95% CI. | |
| --- | --- | --- | --- | --- | --- |
|  |  |  |  | lower | upper |
| Gender | 0.060 | 1.062 | 0.820 | 0.632 | 1.786 |
| Type | -0.788 | 0.455 | 0.034 | 0.220 | 0.940 |
| Age(<20 years) |  |  |  |  |  |
| Age(20-40 years) | 0.133 | 1.143 | 0.788 | 0.431 | 3.028 |
| Age(40-60years) | 0.653 | 1.921 | 0.194 | 0.717 | 5.153 |
| Age(≥60 years) | 1.094 | 2.986 | 0.039 | 1.058 | 8.432 |

Abbreviation: OR: odds ratio; CI: confidence interval

### The Duration of IgM and IgG

Analysis of the 363 serological samples showed that the positive rate of IgG was higher than IgM, and the immune response persistence of IgG was longer than that of IgM antibody, which was consistent with the current research[17]. The median duration of IgM and IgG antibodies was 7 months. We observed duration of both antibodies persisted more than 10 months. It suggested that there may be a long-term immune response after infection with SARS-CoV-2[18]. In order to comprehend the dynamics of antibody response, we took the occurrence of negative antibody as failure, and depicted the survival curve of differential antibodies. As shown in Figure 1, IgM antibody positive rates declined gradually over time after natural infection with SARS-CoV-2. Asymptomatic cases were easier turn out to negative. IgG antibody prevalence declined gradually after the sixth month(Figure 2), with duration last more than 10 months. The total positive rate of symptomatic is higher than that of asymptomatic. As well as the long-term duration. The disappearance time of the two specific antibodies still needs further observation.

**Figure 1.**
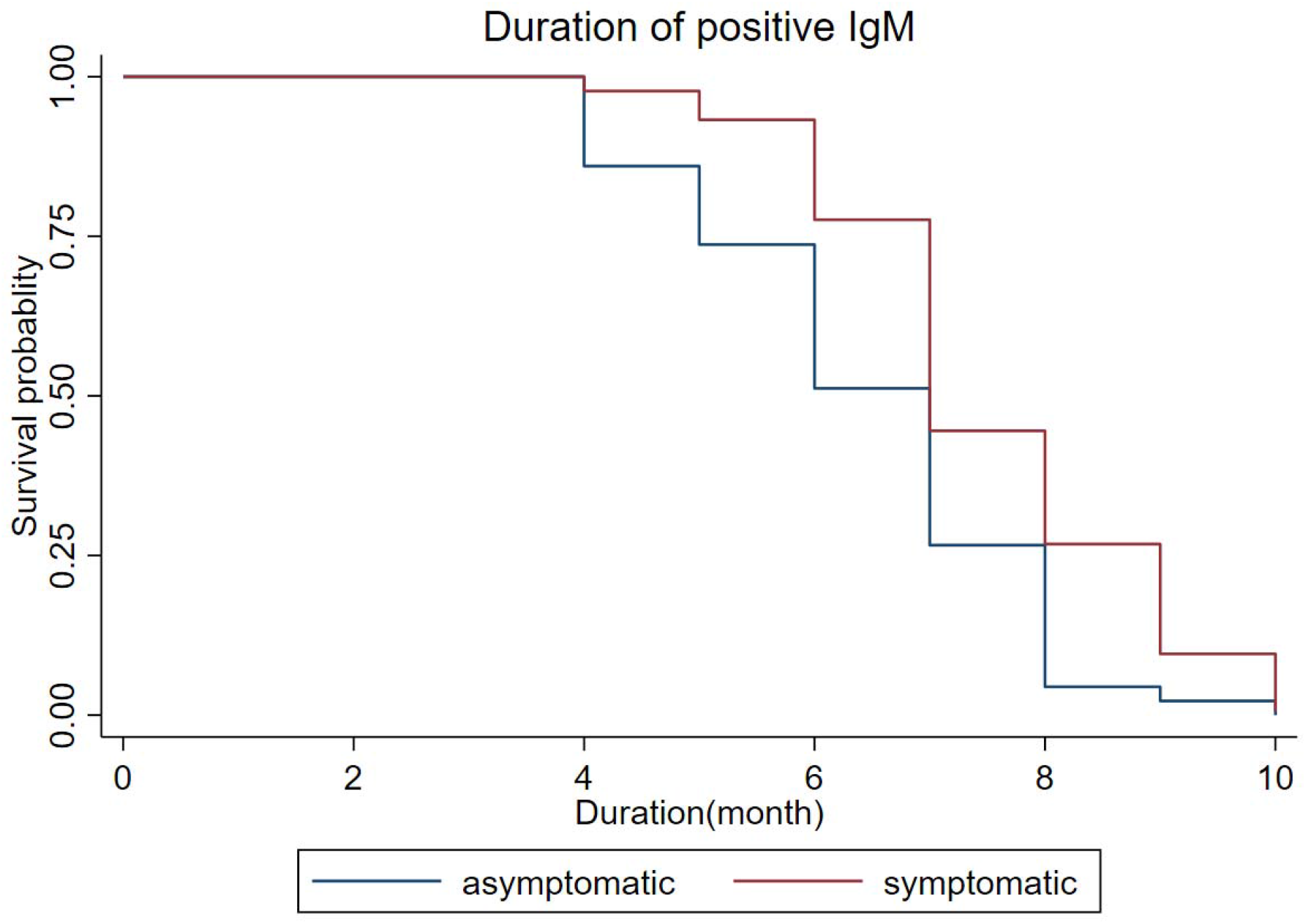
Duration of IgM antibodies among asymptomatic and symptomatic.

**Figure 2.**
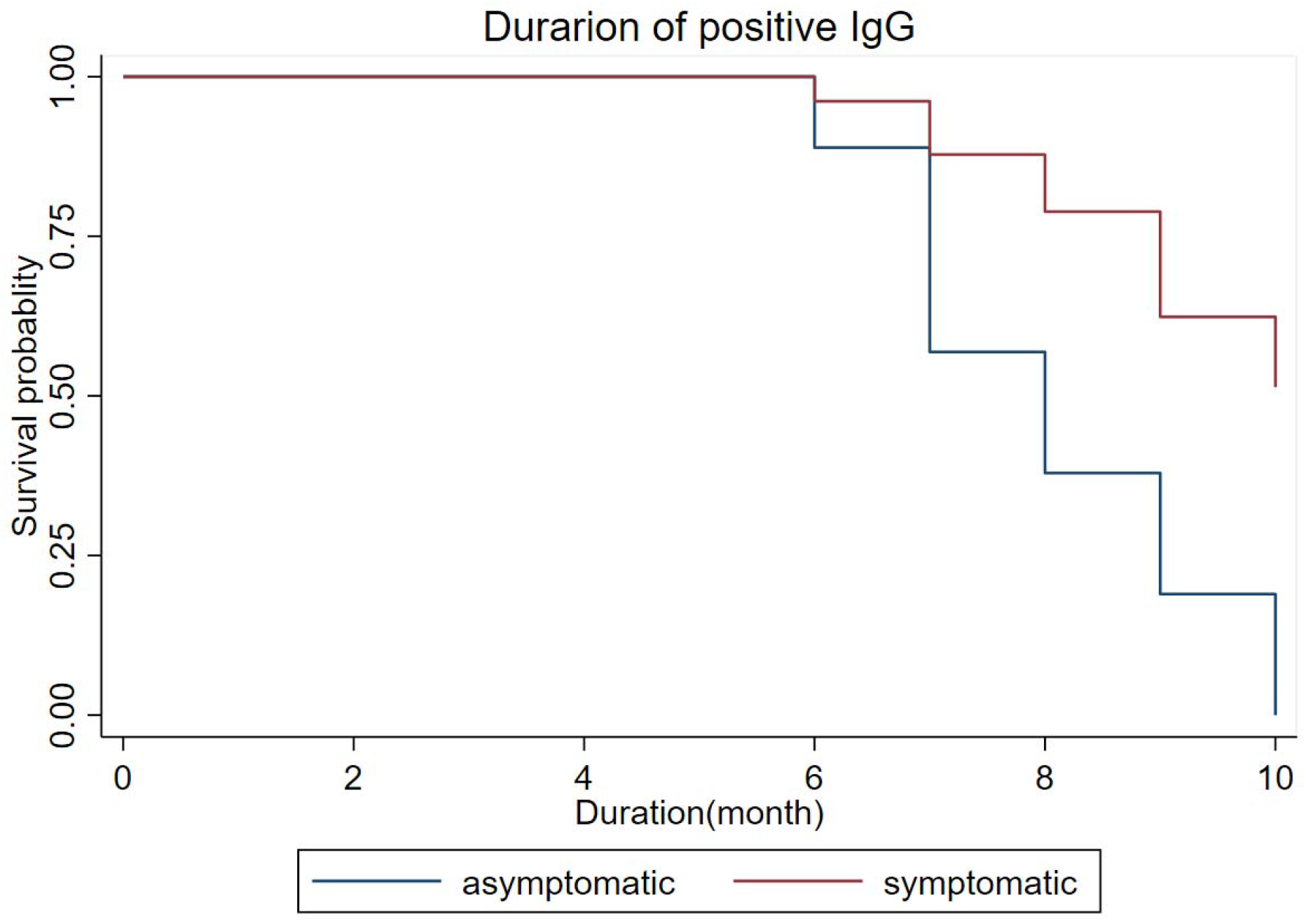
Duration of IgG antibodies among asymptomatic and symptomatic.

## Discussion

Our study contributes more essential information for convalescents about the durablity and stability of antibody response to SARS-CoV-2. Human immune response is usually measured in the blood, and IgG and IgM antibodies are thought as immune memory markers[19]. We analysed the serological outcomes from 177 convalescents of COVID-19 and risk factors. After adjustment for infected type and age, men had more than twice the odds of having IgM antibody positive compared with women (OR=2.419, 95% CI:[1.232, 4.751]). Participants without symptoms were nearly 0.5 times IgG seropositive than those with symptoms (OR=0.455, 95% CI: [0.220, 0.940]). Among different age groups between symptomatic cases and asymptomatic cases with statistical difference(χ^2^= 14.0671,*P*=0.003). People aged ≥ 60 were nearly 3 times IgG seropositive than those who aged < 20 years (OR=2.986, 95% CI: [1.058, 8.432]). Both IgM and IgG seroprevalence declined gradually over time after natural infection with SARS-CoV-2. We also observed seroprevalence of convalescents can persist more than 10 months, the disappearance still needs further observation.

Both IgM and IgG seroprevalence declined gradually over time after natural infection with SARS-CoV-2, with seropositive antibodies lasted more than 10 months. It was demonstrated that response wanes over time of convalescence rather than the sake of repeated donations[20]. Asymptomatic cases were easier turn out to negative. The total positive rate of IgG antibody is higher than that of IgM antibody. The seroprevalence of IgM in infected men was higher than women. Gender differences may affect SARS-CoV-2 immune response outcome[21]. When considering subgroups by gender, women are more likely have higher frequency than in men. After analysis of 363 serological samples collected, it was concluded that the positive rate of IgG was significantly higher than that of IgM, and the immune response persistence of IgG was longer than IgM. Meanwhile, we observed duration of specific antibody was last more than 10 months from our data. Previous study reported was 8 months[14].

Ideally, this study should be based on continuous detection at time points, but in reality it is difficult to carry out continuous systematic sample collection, and not all patients have continuous observation data, which limits the continuous observation of individual antibody response reports. Our data do not yet know whether our individuals sustain low and stable levels after an initial drop in antibody levels, and the inactivation time of specific antibodies generated by natural infection with SARS-CoV-2, which requires further tracking and testing.

Future studies will require large sample sizes of long-term, continuous data on specific immune memory for SARS-CoV-2 to assess its association with protection. They can be conducted with long-term follow-up and detection to investigate factors strongly related to serological levels and antibody dynamics over time, provided deep insight into the immune response to SARS-CoV-2 of convalescents and better guide the development of vaccines and therapeutics.

## Data Availability

The raw/processed data required to reproduce these findings cannot be shared at this time as the data also forms part of an ongoing study.

## Author contributions

CL and LJZ consulted the literature, analyzed the data, wrote the programs, and was a major contributor in writing the manuscript. XPW, HY, TJX and CKX wrote part of the manuscript. MP, CX, HPY, XWC, XYS and XBD collected the data. HWP, TH, HL, DX and WLX detected serum samples, ZP and ZDZ commented on the content of the article. YZ collected part of the data. LJ and XW made constructive comments on the manuscript. TZ contributed significantly to analysis and manuscript preparation.

## Acknowledgments

The authors thank all the colleagues participating in the Sichuan Field Epidemiology Training Program and Standardized Training of Public Health Physicians in Sichuan Province for their contributions to data collection and manuscript review.

## References

[1] Huang C, Huang L, Wang Y, Li X, Ren L, Gu X, et al. 6-month consequences of COVID-19 in patients discharged from hospital: a cohort study. The Lancet. 2021;397:220–32. https://doi.org/10.1016/s0140-6736(20)32656-8

[2] Xiang F, Wang X, He X, Peng Z, Yang B, Zhang J, et al. Antibody Detection and Dynamic Characteristics in Patients with COVID-19. Clin Infect Dis. 2020. https://doi.org/10.1093/cid/ciaa461

[3] Li Z, Yi Y, Luo X, Xiong N, Liu Y, Li S, et al. Development and clinical application of a rapid IgM-IgG combined antibody test for SARS-CoV-2 infection diagnosis. J Med Virol. 2020;92:1518–24. https://doi.org/10.1002/jmv.25727

[4] De Marinis Y, Sunnerhagen T, Bompada P, Blackberg A, Yang R, Svensson J, et al. Serology assessment of antibody response to SARS-CoV-2 in patients with COVID-19 by rapid IgM/IgG antibody test. Infect Ecol Epidemiol. 2020;10:1821513. https://doi.org/10.1080/20008686.2020.1821513

[5] Röltgen K, Powell AE, Wirz OF, Stevens BA, Hogan CA, Najeeb J, et al. Defining the features and duration of antibody responses to SARS-CoV-2 infection associated with disease severity and outcome. Sci Immunol. 2020;5. https://doi.org/10.1126/sciimmunol.abe0240

[6] To KK, Hung IF, Ip JD, Chu AW, Chan WM, Tam AR, et al. COVID-19 re-infection by a phylogenetically distinct SARS-coronavirus-2 strain confirmed by whole genome sequencing. Clin Infect Dis. 2020. https://doi.org/10.1093/cid/ciaa1275

[7] Chow CC, Chang JC, Gerkin RC, Vattikuti S. Global prediction of unreported SARS-CoV2 infection from observed COVID-19 cases. medRxiv : the preprint server for health sciences. 2020. https://doi.org/10.1101/2020.04.29.20083485

[8] Long QX, Liu BZ, Deng HJ, Wu GC, Deng K, Chen YK, et al. Antibody responses to SARS-CoV-2 in patients with COVID-19. Nature medicine. 2020;26:845–8. https://doi.org/10.1038/s41591-020-0897-1

[9] Hou H, Wang T, Zhang B, Luo Y, Mao L, Wang F, et al. Detection of IgM and IgG antibodies in patients with coronavirus disease 2019. Clinical & Translational Immunology. 2020;9. https://doi.org/10.1002/cti2.1136

[10] Li G, Chen X, Xu A. Profile of specific antibodies to the SARS-associated coronavirus. N Engl J Med. 2003;349:508–9. https://doi.org/10.1056/nejm200307313490520

[11] Wu LP, Wang NC, Chang YH, Tian XY, Na DY, Zhang LY, et al. Duration of antibody responses after severe acute respiratory syndrome. Emerging infectious diseases. 2007;13:1562–4. https://doi.org/10.3201/eid1310.070576

[12] Zhou W, Xu X, Chang Z, Wang H, Zhong X, Tong X, et al. The dynamic changes of serum IgM and IgG against SARS-CoV-2 in patients with COVID-19. J Med Virol. 2021;93:924–33. https://doi.org/10.1002/jmv.26353

[13] Poland GA, Ovsyannikova IG, Kennedy RB. SARS-CoV-2 immunity: review and applications to phase 3 vaccine candidates. The Lancet. 2020;396:1595–606. https://doi.org/10.1016/s0140-6736(20)32137-1

[14] Choe PG, Kim KH, Kang CK, Suh HJ, Kang E, Lee SY, et al. Antibody Responses 8 Months after Asymptomatic or Mild SARS-CoV-2 Infection. Emerging infectious diseases. 2020;27. https://doi.org/10.3201/eid2703.204543

[15] Guo X, Zeng L, Huang Z, He Y, Zhang Z, Zhong Z. Longer Duration of SARS-CoV-2 Infection in a Case of Mild COVID-19 With Weak Production of the Specific IgM and IgG Antibodies. Front Immunol. 2020;11:1936. https://doi.org/10.3389/fimmu.2020.01936

[16] Lai CC, Wang JH, Hsueh PR. Population-based seroprevalence surveys of anti-SARS-CoV-2 antibody: An up-to-date review. Int J Infect Dis. 2020. https://doi.org/10.1016/j.ijid.2020.10.011

[17] Wang B, Wang L, Kong X, Geng J, Xiao D, Ma C, et al. Long-term coexistence of SARS-CoV-2 with antibody response in COVID-19 patients. J Med Virol. 2020;92:1684–9. https://doi.org/10.1002/jmv.25946

[18] Bruni M, Cecatiello V, Diaz-Basabe A, Lattanzi G, Mileti E, Monzani S, et al. Persistence of Anti-SARS-CoV-2 Antibodies in Non-Hospitalized COVID-19 Convalescent Health Care Workers. Journal of Clinical Medicine. 2020;9. https://doi.org/10.3390/jcm9103188

[19] Sette A, Crotty S. Adaptive immunity to SARS-CoV-2 and COVID-19. Cell. 2021. https://doi.org/10.1016/j.cell.2021.01.007

[20] Perreault J, Tremblay T, Fournier MJ, Drouin M, Beaudoin-Bussières G, Prévost J, et al. Waning of SARS-CoV-2 RBD antibodies in longitudinal convalescent plasma samples within 4 months after symptom onset. Blood. 2020;136:2588–91. https://doi.org/10.1182/blood.2020008367

[21] Takahashi T, Ellingson MK, Wong P, Israelow B, Lucas C, Klein J, et al. Sex differences in immune responses that underlie COVID-19 disease outcomes. Nature. 2020;588:315–20. https://doi.org/10.1038/s41586-020-2700-3

